# Performance of Google NotebookLM for AI-assisted data extraction and consensus statement generation in a heterogenous systematic review on inflammatory bowel disease, obesity, and cardiometabolic comorbidities: A Methodological Report

**DOI:** 10.64898/2026.06.16.26355773

**Authors:** Sami Samaan, Jalpa Devi, Matthew Vincent, Shannon Coombs, Priya Sehgal, Mouhand Mouhamed, Victoria Rai, Amanda M. Johnson, Andres J. Yarur, Edward L. Barnes, Parakkal Deepak

## Abstract

**Background:** Large language models (LLMs) offer promise for systematic review data extraction, but performance in complex multidisciplinary domains and utility for clinical statement generation remain insufficiently described.

**Objectives:** To evaluate Google NotebookLM for AI-assisted data extraction and RAND/UCLA consensus statement generation in a systematic review of IBD, obesity, and cardiometabolic comorbidities.

**Methods:** Studies were organized into domain-specific notebooks; structured prompts generated standardized evidence tables. Two independent reviewers validated outputs against full-text articles using a four-category error classification. Cell-level accuracy and critical accuracy (cells free of major factual errors) were the primary metrics; workflow time was compared against a published conventional extraction benchmark. Concordance between AI-generated and expert-finalized statements was assessed.

**Results:** Across 57 articles, 1,710 data cells were extracted; 151 (8.83%) were flagged, yielding 91.17% cell-level accuracy. Major factual errors occurred in only 4 cells (0.23%), for a critical accuracy of 99.77%. Most errors were minor omissions (59.6%) or incomplete extractions (30.5%); domain error rates ranged from 7.08% to 11.33%. The pipeline required 17.7 versus a projected 165.1 person-hours (89.3% reduction). PICO-structured prompting generated 70 candidate statements; 58 of 112 finalized panel statements (51.8%) were AI-derived, and 85.7% were retained in the finalized set.

**Conclusion:** Google NotebookLM demonstrates feasibility as a primary extraction and synthesis tool in a multidisciplinary systematic review, with extractive incompleteness as the principal limitation and substantial time savings over conventional approaches. Its novel application to RAND/UCLA consensus statement generation extends AI-assisted evidence synthesis to clinical consensus generation workflow.

## I. Introduction

Systematic reviews provide clinicians with the highest quality of evidence for clinical care, yet data extraction remains one of the most labor-intensive and error-prone steps in the review process. Cochrane guidelines recommend independent extraction by two reviewers to minimize human error [1]. However, this manual approach requires significant labor and resources, particularly for large-scope reviews [2, 3], and concerns have been raised regarding its validity [4].

The rapid advancement of large language models (LLMs) has created opportunities to automate or semi-automate extraction stages of systematic reviews [5, 6]. Tools such as ChatGPT and Claude have demonstrated accuracy ranging from 65% to 97% in extracting data from randomized clinical trials (RCTs), depending on task complexity, prompt quality, and the extent of human involvement [7, 8]. The speed of AI-assisted approach, combined with its high accuracy, put LLM-assisted tools as a promising pipeline for modern systematic review methodology.

However, most published work has focused on quantitative data extraction from RCTs in limited clinical domains. The application of LLM-based extraction to reviews characterized by both disciplinary and methodological heterogeneity, and downstream applications of AI-generated data such as the development of clinical consensus statements, remains insufficiently described. As part of the Modulate Obesity and relateD metabolic compllcations For Yielding improvements in IBD outcomes (MODIFY-IBD) effort [9], we conducted a systematic review of interventional strategies in obesity and cardiometabolic morbidities in patients with IBD.

Here we describe the methodological pipeline employed, in which Google NotebookLM, a retrieval-augmented generative (RAG) AI platform, served as the principal tool for data extraction, followed by manual validation and a novel application to RAND/UCLA Appropriateness Method consensus statement generation. We assess its accuracy and time-saving performance with the aim of providing a reproducible protocol adaptable by other investigators undertaking large-scale systematic reviews in complex clinical domains.

## Methods

### 1. Systematic Review Overview

The systematic review was conducted as per the Preferred Reporting Items for Systematic Reviews and Meta-analyses (PRISMA) 2020 statement [10] with a registered PROSPERO protocol [11], summarizing the effectiveness, safety, weight, and IBD-specific outcomes of cardiometabolic interventions in adults with IBD. Eligible designs included RCTs, cohort, case-control, cross-sectional, and population-based registry studies spanning medical, surgical, diet, and lifestyle interventions.

### 2. Tool Selection and Notebook Organization

Google NotebookLM, operating on the Gemini 3 model, was selected as the primary AI-assisted extraction tool. All data extraction was performed on January 26, 2026. As a RAG platform, NotebookLM confines its generative responses exclusively to user-uploaded source documents, substantially mitigating the risk of hallucination inherent to general-purpose LLMs [12], and supports multi-PDF upload, full-text processing of tables and figures, and in-source citation. A primary notebook held all eligible PDFs; an initial topic-identification prompt defined the main thematic domains, and dedicated secondary notebooks were created for each (GLP-1RA, bariatric procedures, pharmacologic interventions of cardiometabolic conditions, and diet and lifestyle). This architecture enabled domain-targeted prompting and reduced interference from off-topic content **(Supplementary Methods).**

### 3. Extraction Prompt Development and Evidence Table Generation

Structured extraction prompts were developed through an iterative pilot-testing process applied to a subset of ten articles spanning different study designs and subtopics. Initial prompt drafts were designed following the Cochrane Handbook for Systematic Reviews of Interventions recommendations regarding data collection domains [1], and a manually generated data collection sheet. Prompt refinement continued until outputs were consistent, complete, and correctly formatted extraction across ten consecutive pilot articles without modification. The finalized extraction prompt generated a standardized evidence table including 30 predefined variables (**Supplementary Methods**).

Specific instructions within the prompt guided extraction standards, including: reporting any unreported variable as ‘NR’ (not reported); prohibition of generating information not explicitly present in the source document; inclusion of percentage values with corresponding raw counts in parentheses; reporting of outcomes where no statistically significant difference was observed; addition of a supplementary column for relevant information not captured by predefined variables; avoidance of truncating data entries with ellipses or empty fields; and explicit definition of primary and secondary outcomes where applicable. The complete finalized prompt text is provided in **Supplementary Table 1**. Where the source of a data point was ambiguous, supplementary prompts requesting explicit citation of the originating document were applied.

### 4. Manual Validation and Statistical Analysis

All AI-generated evidence tables underwent systematic manual validation by two independent reviewers (SS, JD), who compared each extracted data point against the full text of the corresponding source article and classified each cell using a standardized four-category error taxonomy **(Table 1)**. Discrepancies were resolved by consensus with the senior author (PD). This dual-reviewer, human-in-the-loop step extends single-reviewer LLM-assisted models described in recent literature [7].

**Table 1:** Four-category classification system and definitions for errors in AI-Extracted data.

|  |  |
| --- | --- |
| **Minor Omission (Blue)** | Minor information is missing, where the omission would not affect the interpretation of the extracted data. |
| **Incomplete Extraction (Yellow)** | Incomplete detail, where the extracted information was partially correct but required reviewer supplementation to provide comprehensive interpretation of the extracted data. |
| **Column Misclassification (Orange)** | Correct information placed in an incorrect column. |
| **Major Factual (Red)** | Major error, where the extracted data was incorrect or substantially misrepresented the referenced source. |

Extraction performance was quantified at the cell and study levels. The primary metric, the cell-level error rate, was defined as flagged cells divided by total extracted cells; critical accuracy was the proportion of cells free of major factual errors; and rates were computed separately per subtopic. Study-level error frequency and distribution, error-type proportions, and field-level vulnerability (error counts aggregated by data column) were summarized using descriptive statistics in Microsoft Excel; no inferential statistics were applied.

### 5. RAND/UCLA Consensus Statement Generation and Concordance Analysis

Google NotebookLM was used to generate clinical statements for a RAND/UCLA Appropriateness Method expert consensus process [13]. Statement generation was performed within the primary notebook using a PICO-structured prompt, containing all articles supplemented by the subtopic-specific evidence tables (**Supplementary Methods**).

Statement generation was iterative; outputs were refined by the study team (SS, JD, PD) and RAND/UCLA methodologists (ELB, VR) to ensure clinical precision, evidence-matched strength, and freedom from justification text that could bias expert ratings, then formatted to RAND/UCLA conventions and distributed to a multidisciplinary expert panel for iterative rating rounds (Figure1). To assess statement quality, a paired concordance analysis compared the initial AI-generated list (n = 70) with the finalized expert-reviewed list (n = 112); each finalized statement was classified using a six-category modification taxonomy (**Supplementary Table 2**), with qualitative review of recurrent modification themes.

### 6. Workflow Time Analysis

Workflow time was estimated retrospectively across five defined components (**Supplementary Methods**). Total pipeline person-hours (A+B+C) were derived as: [(AI generation) + (SS validation × 57) + (JD validation × 57) + (consensus)] / 60. A conventional dual-extraction benchmark was calculated by applying the published rate of 86.9 minutes per study per reviewer (Lai et al.[14]) to the present 57-study, dual-reviewer pipeline (165.1 person-hours). Percentage reduction was calculated as (benchmark − pipeline) / benchmark × 100. All estimates are reported as central (typical) values with conservative and upper-bound ranges; no inferential statistics were applied.

### 7. Supplementary Analytical Prompts

A set of supplementary analytical prompts was applied to the primary and secondary notebooks to deepen the understanding of the evidence base and identify unstated assumptions, evidence gaps, and inter-study contradictions. These prompts included requests for identification of clinically groundbreaking insights, adversarial review of methodological flaws, mapping of unstated assumptions, generation of unanswered expert-level questions, and systematic identification of contradictions between sources (**Supplementary Table 3**).

## II. Results

### 1. Included Studies and Evidence Table Generation

Following systematic search and screening, 57 eligible articles were included (**Figure 2**), spanning multiple study designs, geographic regions, and outcome domains. Subtopic analysis within the primary notebook identified four principal domains, used to structure the secondary notebooks: (1) GLP-1RA; (2) bariatric procedures; (3) pharmacologic interventions of cardiometabolic conditions; and (4) diet and lifestyle interventions. The extraction prompt generated one evidence table per domain, totaling 1,710 cells: bariatric procedures (n = 24; 720 cells), GLP-1RA (n = 15; 450 cells), pharmacologic cardiometabolic interventions (n = 7; 210 cells), and diet and lifestyle (n = 11; 330 cells), with each study yielding 30 extractable data cells.

**Figure 1:**
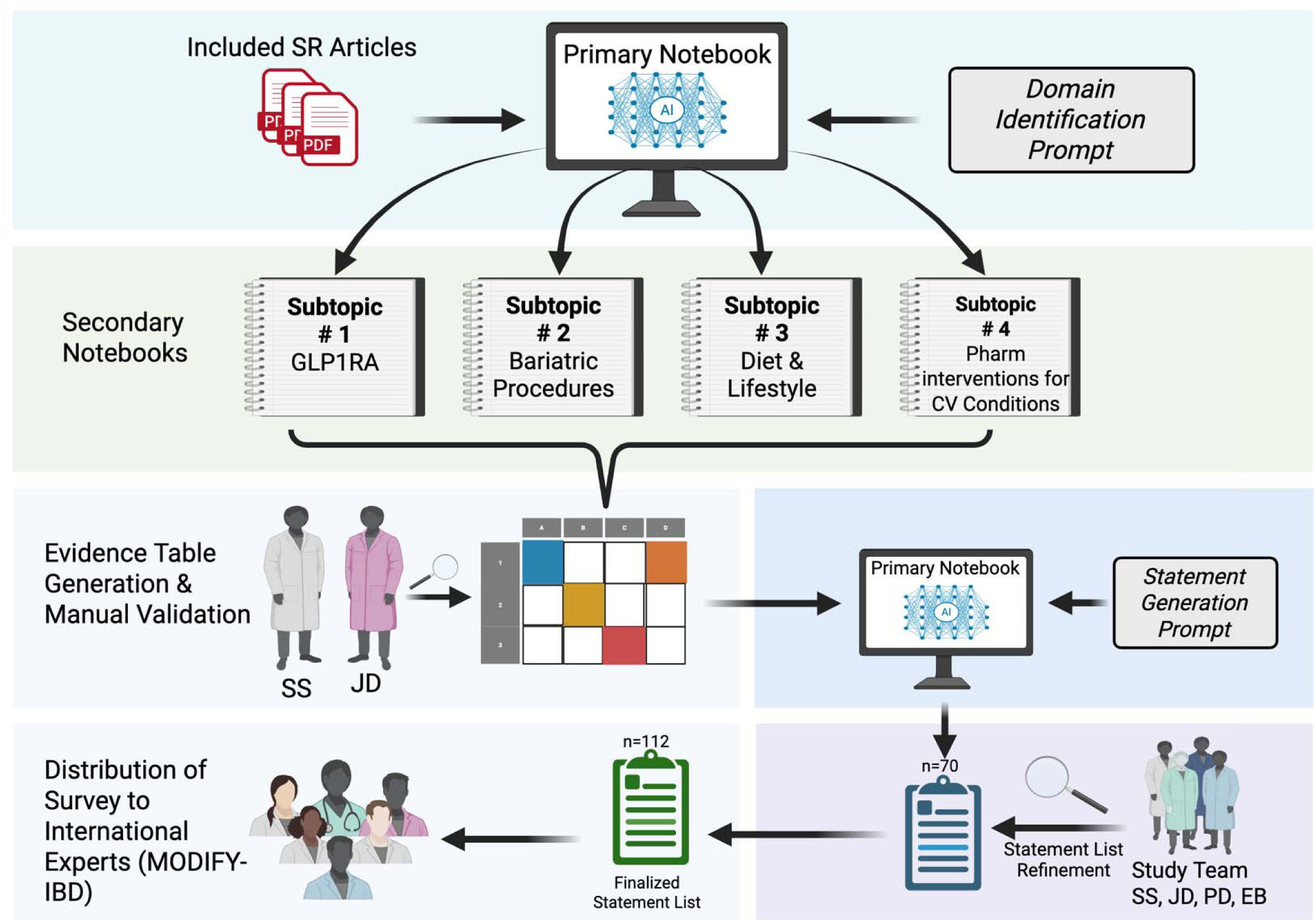
Workflow of NotebookLM-assisted pipeline for data extraction and consensus statements generation. *Abbreviations*: CV, Cardiovascular; GLP1RA, Glucagon-Like Peptide-1 Receptor Agonist; MODIFY-IBD, Modulate and relateD metabolic complications For Yielding improvements in IBD outcomes; Pharm, Pharmacological ments te Obesity

**Figure 2:**
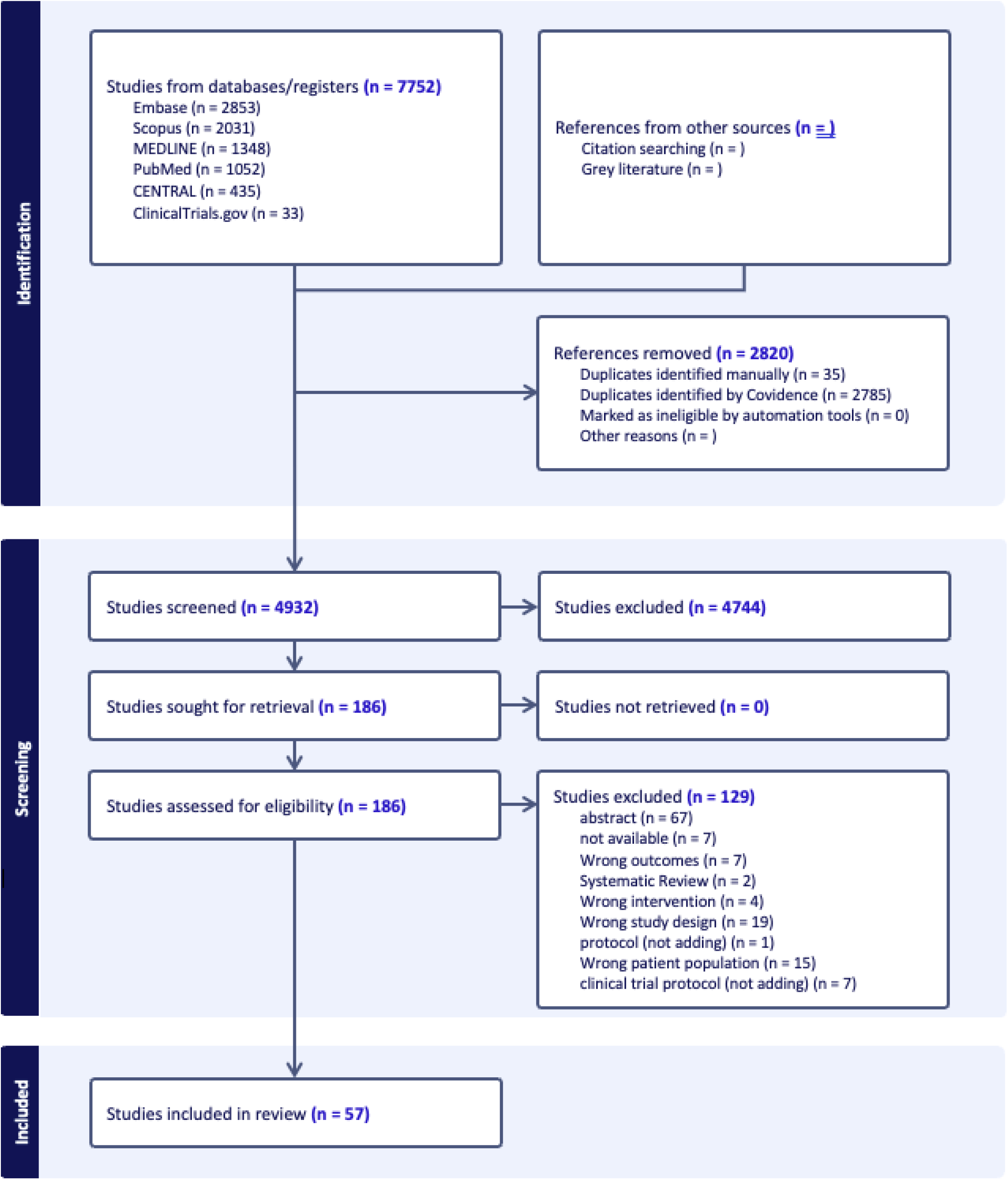
PRISMA 2020 study selection flow diagram for the parent systematic review (PROSPERO CRD420251174653)

### 2. Overall Extraction Accuracy

Of the 1,710 cells reviewed, 151 (8.83%) were flagged as containing at least one extraction error, yielding a cell-level accuracy of 91.17%. Major factual errors affected only 4 cells across 3 studies (0.23% of all cells), for a critical accuracy of 99.77%. At the study level, 3 of 57 studies (5.3%) were extracted with zero errors; the remaining 54 (94.7%) contained at least one flagged cell, with a mean of 2.65 errors per study (median 3; SD 1.33; range 0–5). The distribution was right-skewed, with most studies (n = 42; 73.7%) exhibiting three or fewer errors (**Supplementary Figure 1**). The predominant error type was minor omission (n = 90; 59.6%), followed by incomplete extraction (n = 46; 30.5%), column misclassification (n = 11; 7.3%), and major factual error (n = 4; 2.6%) (**Table 2**).

**Table 2:** Manual validation results, by subtopic domain.

| Domain | Studies (n) | Blue | Red | Yellow | Orange | Total errors | Error rate (%) |
| --- | --- | --- | --- | --- | --- | --- | --- |
| Bariatric Procedures | 24 | 30 | 2 | 15 | 4 | 51 | 7.08 |
| GLP-1RA | 15 | 26 | 1 | 19 | 5 | 51 | 11.33 |
| CVD/IBD | 7 | 9 | 0 | 10 | 2 | 21 | 10 |
| Diet & Lifestyle | 11 | 25 | 1 | 2 | 0 | 28 | 8.48 |
| **Total** | 57 | 90 | 4 | 46 | 11 | 151 | 8.83 |

*Blue = minor omission; Red = major factual error; Yellow = incomplete extraction; Orange = column missclassification. Error rate (%) = domain total errors / domain total cells × 100.*

### 3. Subtopic-specific Errors and Field-Level Vulnerability

Error rates and types varied substantially across the four domains (**Figure 3**). The GLP-1RA and pharmacologic cardiometabolic conditions domains exhibited the highest error rates (10.00% and 8.48%, respectively), and bariatric procedures the lowest (7.08%). However, the profile of errors differed markedly between domains, indicating qualitative as well as quantitative variation in AI extraction performance.

**Figure 3:**
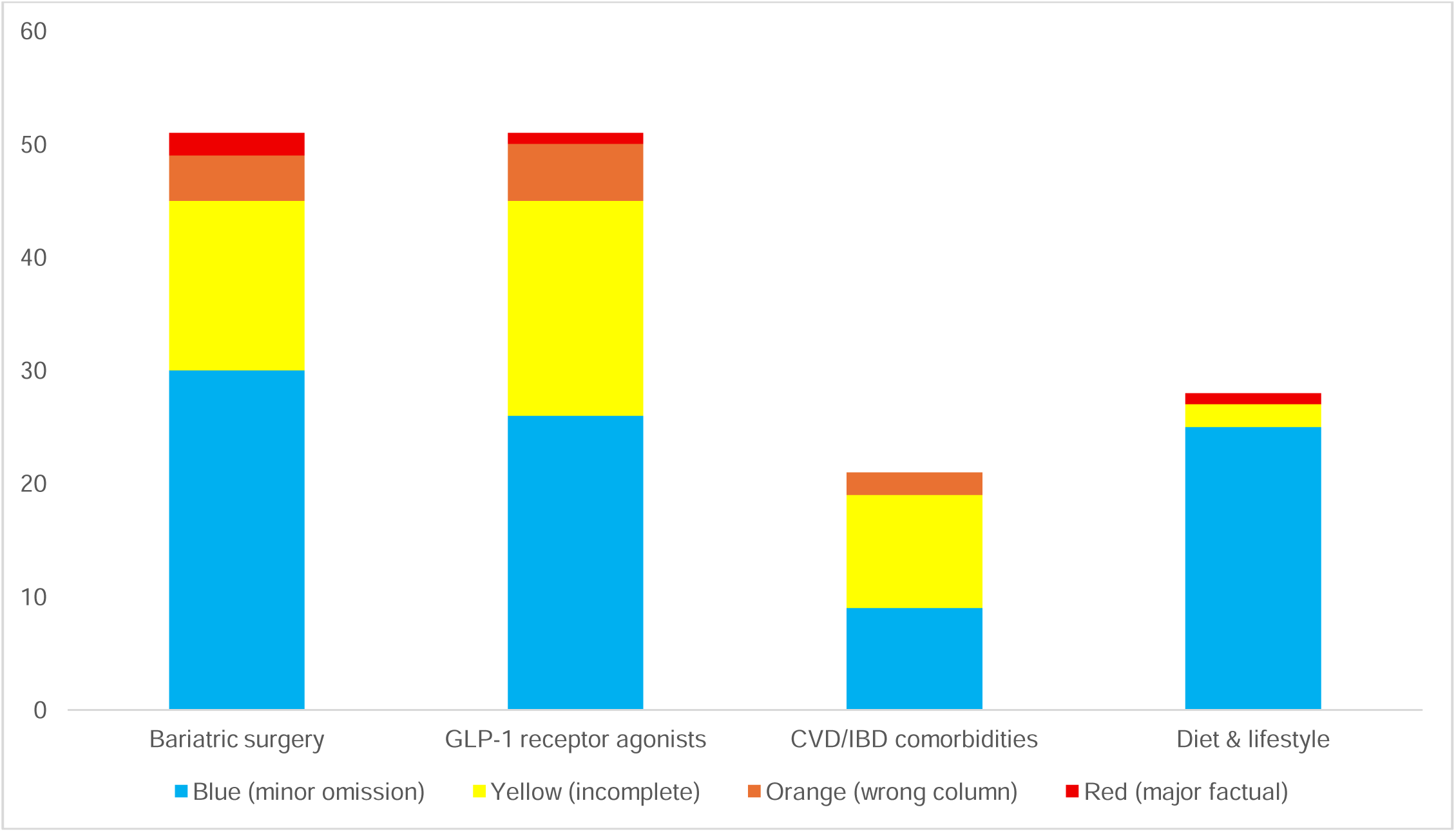
Absolute count and composition of AI extraction errors by subtopic, stratified by error category. *y-axis: absolute count of errors; Blue = minor omission; yellow = incomplete extraction; red = major factual error; orange = correct information placed in wrong column. Numbers above bars indicate total errors per domain.*

In the bariatric procedures domain, errors were predominantly minor omissions (n = 30; 58.8%), and included the greatest number of column misclassifications; GLP-1RA showed a heterogeneous profile (minor omission n = 26, 51.0%; incomplete extraction n = 19, 37.3%; column misclassification n = 5, 9.8%; one major factual error); the pharmacologic domain, despite its small sample (n = 7), was driven by incomplete extraction concentrated in IBD-outcome and subgroup fields; and diet and lifestyle was least heterogeneous, with minor omissions comprising 89.3% of its 28 flagged cells, largely in the Study Title field.

Errors were not uniformly distributed across columns but concentrated in a small number of clinically complex or inconsistently reported fields (**Figure 4**). The IBD outcomes field was the single most error-prone column (30 cells; 19.9% of all errors), consistent with the heterogeneity of IBD outcome reporting, in which variable disease activity indices, composite endpoints, and non-standardized nomenclature collectively increase the interpretive burden placed on the extraction model.

**Figure 4:**
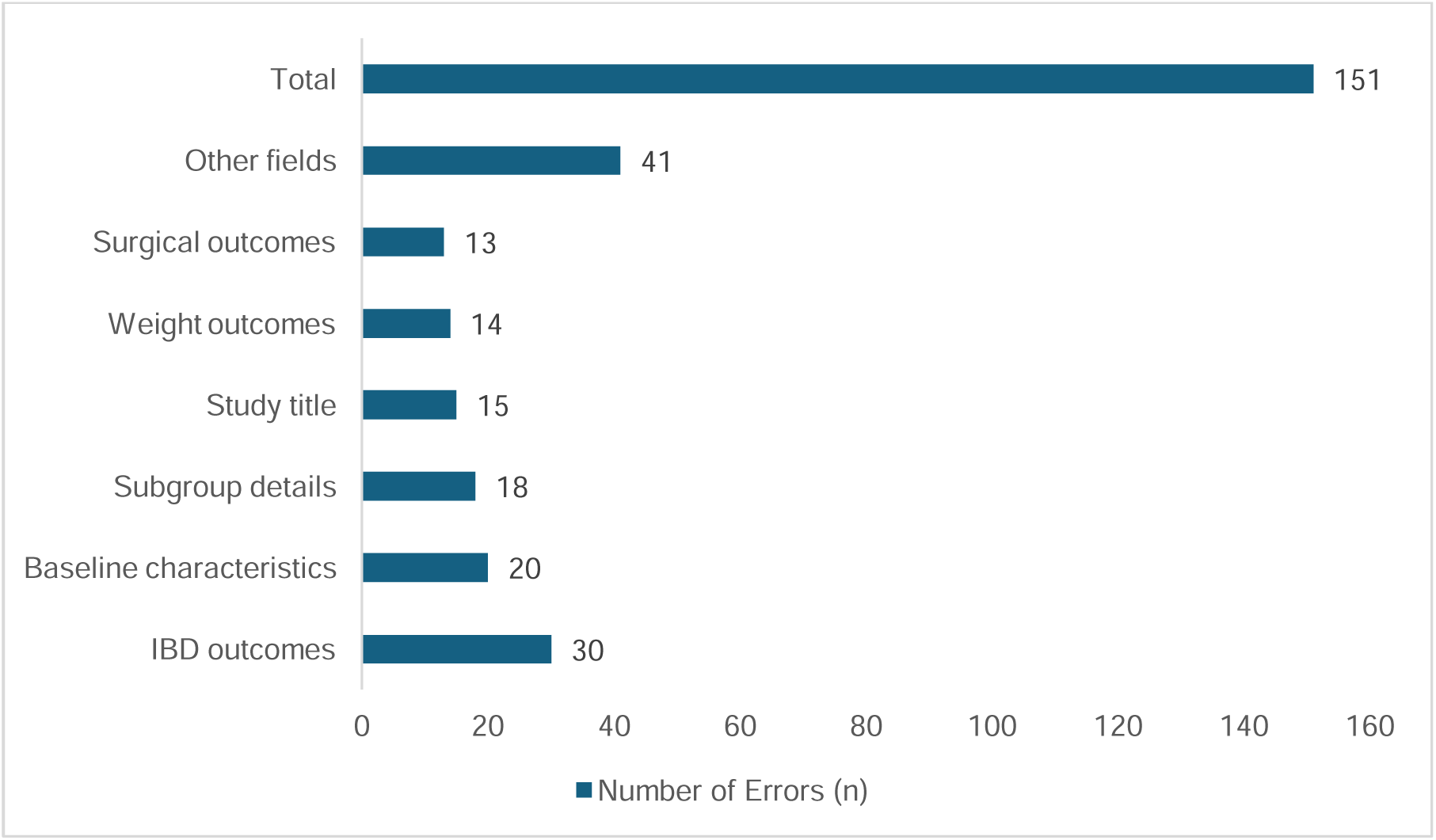
Distribution of AI extraction errors by data field. *Fields ranked in descending order of total errors. Only fields with at least one flagged cell are displayed. IBD = inflammatory bowel disease; CV = cardiovascular.*

Baseline characteristics ranked second (n = 20; 13.2%; predominantly minor omissions), subgroup details third (n = 18; 11.9%; predominantly incomplete extraction), followed by study title (n = 15; 9.9%; diet and lifestyle formatting artefacts), weight outcomes (n = 14; 9.3%), and bariatric procedural outcomes (n = 13; 8.6%). Collectively, the six highest-error fields accounted for 110 of 151 errors (72.8%) (**Supplementary Table 4**).

### 4. Statement Concordance Analysis

The AI-generated list of 70 clinical statements expanded to a finalized list of 112, a net increase of 42 following expert review (**Table 3**). Of the 112 finalized statements, 58 (51.8%) were derived from the AI-list and 54 (48.2%) were added de novo. Among the AI-list, 29 (25.9%) were accepted verbatim or near-verbatim, 17 (15.2%) underwent minor edits, 5 (4.5%) required substantial refinement, and 7 (6.2%) resulted from structural split or combination. Overall, 53 (47.3%) were clinically acceptable without major revision. Sixty of the 70 AI-generated statements (85.7%) contributed to at least one finalized statement, while 10 (14.3%) were deleted, addressing topics such as prognostic biomarkers, microbiome-related outcomes, perioperative metabolic optimization, and de novo IBD risk following bariatric surgery, reflecting insufficient evidence or non-actionability in the RAND/UCLA format. The 54 de novo statements addressed questions beyond the original review scope, mainly in pharmacotherapy appropriateness (statin initiation, cardiovascular risk modification, comparative anti-obesity agents), endobariatric procedures, and dietary interventions, largely relating to weight management in the general population. Qualitative review of modified statements identified five recurrent expert editing patterns: evidence-strength adjustment (e.g., changing “should be considered” to “is appropriate”), population expansion (from “obesity” to “obesity or overweight”), terminology standardization, splitting composite statements into individually ratable sub-items, and clinical-scope redefinition (**Supplementary Table 5**).

**Table 3.** Statement-level concordance analysis: AI-generated list versus expert-refined consensus statements.

| Category | n | % of final survey | Description |
| --- | --- | --- | --- |
| Verbatim/Near-verbatim | 29 | 25.9 | Clinical meaning and structure preserved |
| Minor linguistic edit | 17 | 15.2 | Terminology, abbreviation, spelling, or scope; core recommendation unchanged |
| Substantial clinical refinement | 5 | 4.5 | Qualifying language, recommendation direction, or clinical scope modified |
| Split/Combined | 7 | 6.2 | Disaggregated into sub-items or merged |
| De novo expert addition | 54 | 48.2 | No AI-generated precursor identified |
| Total | 112 | 100.0 |  |

### 5. Workflow Efficiency

AI table generation (Component A) required 5 minutes in total across all 57 studies, reflecting the source-level processing architecture of NotebookLM. Human validation (Component B) required a typical 8 minutes per study (SS; range 5–10) and 10 minutes per study (JD; range 8–20), a combined burden of 1,026 minutes (17.1 person-hours); consensus resolution (Component C) required 30 minutes. Total pipeline time was 17.7 person-hours (conservative 12.9; upper bound 29.1), or 19.7 person-hours including prompt development, with human validation accounting for 96.7% of the total (Table 4). Relative to the projected conventional dual-extraction burden of 165.1 person-hours, the pipeline represents an estimated reduction of 147.4 person-hours (89.3%). For statement generation, AI produced 70 candidate statements in 3 minutes; per-statement human review (70 minutes) and expert iterative refinement (3 hours) totaled 4.22 person-hours (upper bound 6.55), with expert refinement constituting 71% of this total (**Table 4**).

**Table 4.** Retrospective Time Estimates for the AI-Assisted Workflow.

| Component | Responsible | Typical estimate | Total (57 studies) | Person-hours |
| --- | --- | --- | --- | --- |
| A — AI generation | SS only | 5 min (project total) | 5 min | 0.08 |
| B — Validation (SS) | SS | 8 min/study | 456 min | 7.6 |
| B — Validation (JD) | JD | 10 min/study | 570 min | 9.5 |
| C — Consensus resolution | Joint | 30 min (project total) | 30 min | 0.5 |
| A+B+C Total |  |  | 1,061 min | 17.7 |
| D — Prompt development | SS only | 2 hrs. (project total) | — | 2.0 |
| A+B+C+D Total |  |  | — | 19.7 |
| E1 — Statement AI generation | SS only | 3 min (project total; 70 statements) | 3 min | 0.05 |
| E2 — Statement human review | SS, JD, PD | 1 min/statement | 70 min | 1.17 |
| E3 — Expert refinement | SS, JD, PD | 3 hrs. (project total) | — | 3.0 |
| Statement pipeline total |  |  | — | 4.22 |

## III. Discussion

This report describes the design, implementation, and validation of an AI-assisted data extraction and consensus statement generation pipeline using Google NotebookLM, applied to a complex systematic review and consensus statement generation in a GI disease-state examining the multidisciplinary management of IBD, obesity, and cardiometabolic comorbidities. The principal contributions of this work are: (1) a detailed, replicable methodology for the application of a RAG LLM platform to systematic review evidence synthesis across heterogenous clinical domains; (2) a prospective characterization of extraction error type and distribution using a four-category classification system across all extracted cells; and (3) a novel downstream application of AI-assisted evidence synthesis to RAND/UCLA Appropriateness Method consensus statement generation.

### 1. RAG Architecture and Hallucination Mitigation

The RAG architecture of NotebookLM distinguishes it from general-purpose LLMs operating in open-generative mode: it generates responses exclusively from uploaded sources with inline citations, reducing the risk of hallucination that has been identified as a barrier to autonomous AI-driven evidence synthesis [5, 6]. The theoretical basis is well established, with retrieval-augmented models outperforming parametric-only models on knowledge-intensive tasks [15], and a published meta-analysis confirming an accuracy advantage over standard LLMs (pooled OR 1.35, 95% CI 1.19–1.53) [16]. Our cell-level accuracy of 91.2% is modestly below the 95.1–97.9% reported by Lai et al. [14], and exceeds the 77.2% concordance reported by Gartlehner et al. [7]. The difference is attributable to the greater clinical and methodological heterogeneity of our evidence base, which spanned retrospective cohorts, population-based registries, RCTs, and case series across four domains, compared with the predominantly RCT data of prior studies. Peng et al. similarly reported 65.7–71.5% accuracy for Claude 3.5 in sleep medicine [8]. NotebookLM thus achieves accuracy comparable to single-LLM approaches while adding the safeguard of source-constrained generation in a complex multidisciplinary review and expanding the scope to statement generations.

### 2. Error Type Distribution and Prompt Engineering

The predominance of incompleteness-type over factual errors indicates that the principal limitation of the pipeline is insufficient detail extraction rather than generation of incorrect information, a deficit correctable at the human verification step and less detrimental to evidence synthesis. Column misclassification (11 cells across 8 studies), a category absent from prior single-domain validations [7, 8] represents an under-characterized failure mode in complex multi-column tables, concentrated where column domains overlap (e.g., baseline characteristics versus baseline cardiovascular comorbidities); future implementations should address this through column-level micro-definitions and explicit content examples in the extraction prompt. Field-level vulnerability followed a predictable pattern, with IBD outcomes, subgroup details, and weight outcomes, the domains of greatest reporting heterogeneity, accounting for 41.1% of errors, consistent with the outcome-type variation reported by Peng et al. [8]. The multi-notebook, domain-specific workflow minimized off-topic interference and allowed domain-specific terminology in each prompt, consistent with modular approaches identified as an emerging methodological tool [6]. while iterative prompt refinement, central to output quality [17], aligned with the multiple-sentence prompting approach shown by Peng et al. to improve accuracy by up to 11% [8].

### 3. RAND/UCLA Consensus Statement Generation

To our knowledge, the application of NotebookLM to RAND/UCLA consensus panel statement generation represents a novel use of AI-assisted evidence synthesis in a formal consensus statement development, a setting that requires unambiguous, clinically actionable statements appropriately evaluated to evidence strength [13]. Most AI-generated statements required no substantive revision before panel distribution. The substantial de novo expert contribution (48.2%) should be interpreted in the context of pipeline design rather than as an AI limitation: the AI list was generated from a defined evidence body, whereas the expert team drew on broader clinical knowledge and on appropriateness questions relevant to general-population weight management not captured in the IBD-focused review. Future work could evaluate AI rating of clinical statements against expert ratings, as in Sarvari et al., which found substantial concordance between AI outputs and expert-derived recommendations across obesity-management statements[18]. The supplementary analytical prompts further enriched the team’s critical appraisal of the evidence.

### 4. Workflow Efficiency

The estimated 89.3% reduction in person-hours relative to projected conventional dual extraction modestly exceeds the 83.1% reported by Lai et al. [14], in a relatively more homogenous evidence base. Notably, human validation constituted the overwhelming majority of total pipeline time across both extraction and statement generation tasks, suggesting that efficiency gains are concentrated in high-volume primary extraction rather than in reduced human oversight, consistent with the pipeline’s intended role as an accelerant for, rather than substitute for, expert judgement.

### 5. Limitations

Several limitations warrant acknowledgement. First, the absence of a prospectively designated gold-standard subset means the 91.17% accuracy estimate reflects agreement between AI output and post-correction human consensus rather than an independent reference standard; future implementations should designate a blinded validation subset. Second, error rates varied across domains, indicating sensitivity to domain-specific reporting heterogeneity and article complexity. Third, use of a single platform limits comparability; head-to-head evaluations against other RAG-capable tools (e.g., Elicit, Consensus, or Rayyan AI) are warranted. Fourth, time savings were quantified retrospectively; prospective time-on-task measurement using a study-within-reviews framework remains an important priority.

## IV. Conclusion

This study demonstrates the feasibility of Google NotebookLM as a primary tool for data extraction and consensus statement generation in a large-scale, multi-domain systematic review on IBD, obesity, and cardiometabolic comorbidities. The high critical accuracy, the predominance of incompleteness- over factual-error types, and the substantial time savings relative to conventional dual extraction collectively indicate that the principal bottleneck of this pipeline is extractive incompleteness, a deficit that can be fixed through the human-in-the-loop verification step employed.

This report provides an efficient, structured, and replicable framework that may be adapted by other investigators undertaking systematic reviews in complex or multidisciplinary clinical domains. The novel downstream application of this pipeline to RAND/UCLA Appropriateness Method consensus statement generation further expands the potential scope of AI-assisted evidence synthesis beyond data extraction to include the development of clinical guidance.

## Ethics Statement

No human subjects, patient data, and informed consent were collected. Institutional review board approval and informed consent were not required.

## Authors’ contributions

SS conceived the study concept and led the overall design, coordination, and drafting of the manuscript. **JD** contributed to the data validation as the second reviewer. ELB and VR provided methodological expertise for the RAND/UCLA consensus framework, and critically revised the manuscript for important intellectual content. MV, SC, PS, MM, AMJ and **AJY** critically revised the manuscript for important intellectual content. PD conceived the study concept, provided overall supervision of the study, and initial drafting and critical revisions of the manuscript. All authors reviewed, edited, and approved the final version of the protocol

## AI Usage statement

Google NotebookLM was used as the primary AI tool under evaluation for data extraction and consensus statement generation, as described in the Methods. Claude assisted with manuscript editing. All AI-generated outputs were reviewed and verified by the authors, who take full responsibility for the accuracy and integrity of the published work.

## Data availability statement

All data relevant to the study are included in the article and its supplementary materials.

## Abbreviations

AI: Artificial Intelligence
BMI: Body Mass Index
CD: Crohn’s Disease
CVD: Cardiovascular Disease
GLP-1RA: Glucagon-Like Peptide-1 Receptor Agonist
IBD: Inflammatory Bowel Disease
LLM: Large Language Model
MACE: Major Adverse Cardiovascular Event
MASLD: Metabolic Dysfunction-Associated Steatotic Liver Disease
MASH: Metabolic Dysfunction-Associated Steatohepatitis
MD: Mean Difference
MODIFY-IBD: Modulate Obesity and relateD metabolic complications For Yielding improvements in IBD outcomes
NAFLD: Non-Alcoholic Fatty Liver Disease
PICO: Population, Intervention, Comparator, Outcome
PRISMA: Preferred Reporting Items for Systematic Reviews and Meta-Analyses
PROSPERO: International Prospective Register of Systematic Reviews
RCT: Randomized Controlled Trial
SR: Systematic Review
T2DM: Type 2 Diabetes Mellitus
UC: Ulcerative Colitis
VAT: Visceral Adipose Tissue

