## Supplementary Methods for "Performance of Google NotebookLM for AI-assisted data extraction and consensus statement generation in a heterogenous systematic review on inflammatory bowel disease, obesity, and cardiometabolic comorbidities: A Methodological Report"

**S1. Notebook Organization**

A primary notebook was created on the Google NotebookLM platform, and all eligible PDF articles were uploaded as source documents. This primary notebook served as the repository for the complete article set and was subsequently used both for thematic subtopic identification and for consensus statement generation. To facilitate subtopic organization and optimize extraction efficiency, an initial topic-identification prompt was applied to determine the main thematic domains within the source literature. An adapted version of the following prompt was used:

*“Analyze these sources and identify the main thematic topics. Create a table with one row for each theme, then divide the sources between these topics.”*

Based on the AI-generated subtopic table, dedicated secondary notebooks were created for each domain (GLP-1RA, bariatric procedures, pharmacologic interventions of cardiometabolic conditions, and diet and lifestyle), and the corresponding articles were uploaded to each. This architecture enabled targeted prompting within each domain, reducing interference from off-topic content and improving extraction precision. We considered an alternative single-notebook approach in which individual sources are selectively activated or deactivated; however, subtopic-specific secondary notebooks were adopted as the primary workflow to minimize the cognitive overhead of manual source management and to enhance prompt tailoring to each domain.

**S2. Evidence Table Generation and Validation**

The AI-generated evidence tables were saved as notes within the respective subtopic notebooks and subsequently exported to Microsoft Excel format (Microsoft Corporation, Redmond, WA, USA) for manual validation. Exported tables were transferred to a shared cloud-based storage (Microsoft OneDrive) to facilitate collaborative review access. The extraction process was performed iteratively across each subtopic notebook, providing domain-specific evidence tables that summarized the data for the systematic review.

**S3. Domains of variables included in the AI-generated evidence tables**

| **Domain** | **Variables included** |
| --- | --- |
| Study identification | Author(s), publication year, article title, journal name, geographic region of the study population |
| Study design characteristics | Study aim, study type (retrospective cohort, randomized controlled trial, prospective cohort, population-based registry analysis, or other), and total study duration |
| Population characteristics | Baseline characteristics of the study sample, baseline medication exposure (particularly IBD-directed therapies), and baseline cardiovascular comorbidities (hypertension, dyslipidemia, type 2 diabetes mellitus, metabolic syndrome, atherosclerotic cardiovascular disease, and Metabolic Dysfunction-Associated Steatotic Liver Disease (MASLD)/ Non-alcoholic Fatty Liver Disease(NAFLD ). |
| Methodological parameters | Sample size, inclusion criteria, and exclusion criteria |
| Intervention details | Primary intervention and comparator (where applicable) |
| Outcomes | IBD-related outcomes, surgical outcomes (perioperative and postoperative), cardiovascular and metabolic outcomes (blood pressure, glycemic control, lipid profile, MASLD-related outcomes, and MACE), and weight-related outcomes |
| Quantitative data | Weight measure employed (BMI, visceral adipose tissue, waist circumference, or other), effect point estimate, effect measure type (odds ratio, relative risk, mean difference, or other), and duration of follow-up |
| Additional study details | Adverse events, availability and details of subgroup analyses, study limitations, and funding sources and conflicts of interest |

**S4. Generation of RAND/UCLA Expert Consensus Panel Statements:**

The RAND/UCLA method is a well-validated, structured approach to synthesizing scientific evidence with expert clinical judgement, mainly used when high-quality evidence is limited, heterogenous, or not directly applicable to practice. Statements must be designed to be clear, clinically relevant, and ratable by expert panelists on a predetermined appropriateness scale.

PICO-Structured Prompt Used

*“Generate a written summary of the evidence; the aim is to provide information needed for experts in the gastroenterology and obesity field to vote on a set of clinical statements in a RAND/UCLA panel.*

*Please include the referenced studies in-text. Use the PICO (population, intervention, comparison, outcomes) logic to think about the statements, without explicitly mentioning PICO elements.*

*Statements should not use a question format, should not include the reasoning or evidence for the statement, and should use qualifying language (e.g., 'may be appropriate' or 'is appropriate') when the evidence is limited.*

*Statements should follow the structural model: 'In patients with IBD, clinicians should routinely assess Body Mass Index (BMI) as part of standard clinical evaluation.’”*

**S5. Workflow Time Analysis:**

Workflow time was estimated retrospectively using a component-based approach comprising five components:

- **Component A — AI evidence-table generation**: wall-clock time required for NotebookLM to generate the domain evidence tables following prompt submission. Because generation operates at the source level rather than per study, this component reflects total processing time across all studies within a domain.
- **Component B — Human validation**: per-study time required by each independent reviewer (SS, JD) to compare every extracted cell against the corresponding source full text and apply the four-category error classification.
- **Component C — Consensus resolution**: time required to resolve inter-reviewer discrepancies through consensus discussion with the senior author (PD), summed across the four domain sessions.
- **Component D — Prompt development**: one-time investment in iterative pilot prompt engineering, reported separately because it is amortized across the full review and across future applications.
- **Component E — Statement generation**: AI generation of candidate statements, per-statement human review, and expert iterative refinement of the complete statement set, analyzed separately from the extraction pipeline.
