## Supplementary Material for "Performance of Google NotebookLM for AI-assisted data extraction and consensus statement generation in a heterogenous systematic review on inflammatory bowel disease, obesity, and cardiometabolic comorbidities: A Methodological Report"

**Supplementary Table 1:** Finalized data extraction prompt

*As part of data collection for a systematic review, generate an "Evidence Table" where you summarize every single study. I expect to see {# of articles} rows.*

*< variables needed, #30 columns total>*

*author; date; study title; journal name; region; study aim; study type (retrospective, RCT, prospective, population-based, other); study period (total study duration);  baseline characteristics of the sample; baseline medication exposure; Baseline cardiovascular comorbidities ( Hypertension, Dyslipidemia, T2DM, metabolic syndrome; Atherosclerotic CVD; MASLD/NAFLD);  sample size; inclusion criteria; exclusion criteria; primary intervention; comparator; IBD outcomes; Surgical outcomes (anything peri-operative or post-operative); Cardiovascular comorbidities outcomes  (Blood pressure, Glycemic control, Lipid profile, MASLD outcomes, MACE); Weight outcomes; weight measure used (BMI, VAT, waist circumference; other); effect point estimate; effect point type (OR, RR, Mean, etc); adverse events; duration of follow up; subgroup availability (Y/N); subgroup details; limitations; funding/COI*

*<Missing information> 
report it as NR aka Not reported. Never generate information not reported in the sources*

*<Relevant important information that does not fit in the columns above>*

*< If you use words like “higher” “lower” “less/more common” or any words indicating comparison, make sure you list the associated numbers with that >*

*< When including percentages, include counts between parenthesis >*

*< Define primary outcomes and secondary outcomes >*

*< Include outcomes when there is no difference. >*

*< Add a new column for subgroup analysis details   >*

*< Do NOT use “etc” >*

*< Do NOT cut information by adding dots or empty spaces”*

*Add it in an extra column labeled “Extra relevant info”*

*<sanity check> 
Double check each entry against the source to make sure you are not hallucinating data*

**Supplementary Table 2:** Taxonomy of Errors for Clinical Statements Concordance Analysis

| **Category** | **Definition** |
| --- | --- |
| verbatim or near-verbatim retention | Preservation of both clinical meaning and statement structure with only negligible formatting changes |
| Minor edit | Changes in terminology, abbreviation, spelling, or scope that did not alter the core clinical recommendation |
| Substantial refinement | Modification of qualifying language, recommendation direction, or clinical scope |
| Split or combined | A structural rearrangement in which one AI-generated statement was disaggregated into multiple individually ratable sub-items, or conversely, multiple AI-generated statements addressing the same clinical question across disease subtypes (e.g., Crohn’s disease (CD) and ulcerative colitis (UC)) were merged into a single IBD-level statement |
| De novo expert addition | A finalized statement with no identifiable AI-generated precursor |
| Deletions | AI-generated statements absent from the finalized list |

**Supplementary Table 3**: Supplementary Prompts

[[ You are an IBD and Obesity expert with 15 years of experience. Analyze these sources and identify the 3 core insights that practitioners in this field would immediately recognize as groundbreaking. For each insight, explain why it matters and what conventional wisdom it challenges.]]

[[ Act as a harsh peer reviewer. Identify every methodological flaw, logical gap, overclaim, and unsupported leap in these sources. For each weakness, suggest what additional evidence would be needed to strengthen the argument.]]

[[ Create a comprehensive framework that integrates all concepts from these sources. Include: key components, relationships between components, decision trees for application, and edge cases where the framework breaks down.]]

[[ Identify every unstated assumption in these sources. For each assumption, rate how critical it is (1-10) and how likely it is to be wrong. Explain what would change if that assumption were false.]]

[[Based on these sources, generate 15 questions that an expert would ask but that these sources DON'T answer. Prioritize questions that would advance the field or reveal critical gaps in current understanding.]]

[[Extract every actionable step, tool, framework, and technique mentioned across all sources. Organize them into a step-by-step implementation plan with prerequisites, expected outcomes, and potential pitfalls for each step.]]

[[Compare these sources and identify every point where they contradict each other. For each contradiction, explain which source has stronger evidence and why. If both are credible, explain what factors might explain the disagreement.]]

**Supplementary Table 4:** Error counts per category by study

| **Supplementary Table 4: Error Counts per Category by Study** | | | | | | |
| --- | --- | --- | --- | --- | --- | --- |
| **First Author** | **Minor omission** | **Major factual** | **Incomplete extraction** | **Column missclassification** | **Total Errors** | **Error Rate (%)** |
| **── Bariatric Procedures ──** | | | | | | |
| Aelfers | 3 | 0 | 0 | 0 | 3 | 10.0% |
| Aminian | 0 | 2 | 1 | 0 | 3 | 10.0% |
| Bazerbachi | 2 | 0 | 0 | 0 | 2 | 6.7% |
| Braga Neto | 2 | 0 | 0 | 0 | 2 | 6.7% |
| Colombo | 1 | 0 | 0 | 0 | 1 | 3.3% |
| Corbière | 0 | 0 | 0 | 0 | 0 | 0.0% |
| Gupta et al. | 1 | 0 | 0 | 0 | 1 | 3.3% |
| Heshmati | 2 | 0 | 2 | 0 | 4 | 13.3% |
| Honoré | 0 | 0 | 1 | 0 | 1 | 3.3% |
| Hudson | 1 | 0 | 2 | 0 | 3 | 10.0% |
| Igwe | 1 | 0 | 0 | 0 | 1 | 3.3% |
| Johnson | 1 | 0 | 1 | 0 | 2 | 6.7% |
| Keidar | 1 | 0 | 0 | 0 | 1 | 3.3% |
| Lin | 1 | 0 | 0 | 0 | 1 | 3.3% |
| Litmanovich | 3 | 0 | 0 | 0 | 3 | 10.0% |
| Mabeza | 2 | 0 | 1 | 0 | 3 | 10.0% |
| McKenna | 3 | 0 | 1 | 0 | 4 | 13.3% |
| Reenaers | 1 | 0 | 0 | 0 | 1 | 3.3% |
| Sharma | 1 | 0 | 2 | 1 | 4 | 13.3% |
| Stenberg | 0 | 0 | 2 | 1 | 3 | 10.0% |
| Wallhuss | 0 | 0 | 0 | 2 | 2 | 6.7% |
| Desai et al. | 2 | 0 | 1 | 0 | 3 | 10.0% |
| Khan et al. | 0 | 0 | 0 | 0 | 0 | 0.0% |
| Ungar et al. | 2 | 0 | 1 | 0 | 3 | 10.0% |
| **Subtotal** | **30** | **2** | **15** | **4** | **51** | **7.08%** |
| **── GLP-1 Receptor Agonists ──** | | | | | | |
| Anderson et al. | 1 | 0 | 0 | 1 | 2 | 6.7% |
| Ramos Belinchón et al. | 0 | 1 | 0 | 0 | 1 | 3.3% |
| Gorelik et al. | 2 | 0 | 2 | 0 | 4 | 13.3% |
| Clarke et al. | 2 | 0 | 3 | 0 | 5 | 16.7% |
| Desai et al. (a) | 5 | 0 | 0 | 0 | 5 | 16.7% |
| Desai et al. (b) | 0 | 0 | 2 | 0 | 2 | 6.7% |
| Karacabeyli et al. | 2 | 0 | 0 | 2 | 4 | 13.3% |
| Levine et al. | 3 | 0 | 1 | 0 | 4 | 13.3% |
| Nielsen et al. | 2 | 0 | 2 | 1 | 5 | 16.7% |
| Pham et al. | 1 | 0 | 2 | 1 | 4 | 13.3% |
| Saadeh et al. | 2 | 0 | 1 | 0 | 3 | 10.0% |
| Sehgal et al. | 2 | 0 | 2 | 0 | 4 | 13.3% |
| St-Pierre et al. | 2 | 0 | 1 | 0 | 3 | 10.0% |
| Villumsen et al. | 1 | 0 | 1 | 0 | 2 | 6.7% |
| Weng et al. | 1 | 0 | 2 | 0 | 3 | 10.0% |
| **Subtotal** | **26** | **1** | **19** | **5** | **51** | **11.33%** |
| **── Pharm CV Conditions ──** | | | | | | |
| AlRasheed et al. | 2 | 0 | 0 | 1 | 3 | 10.0% |
| Mantaka et al. | 2 | 0 | 1 | 1 | 4 | 13.3% |
| Jacobs et al. | 0 | 0 | 5 | 0 | 5 | 16.7% |
| Lund et al. | 2 | 0 | 0 | 0 | 2 | 6.7% |
| Patel et al. | 1 | 0 | 2 | 0 | 3 | 10.0% |
| Petrov et al. | 2 | 0 | 0 | 0 | 2 | 6.7% |
| Zakroysky et al. | 0 | 0 | 2 | 0 | 2 | 6.7% |
| **Subtotal** | **9** | **0** | **10** | **2** | **21** | **10.00%** |
| **── Diet & Lifestyle ──** | | | | | | |
| Chicco et al. | 2 | 0 | 0 | 0 | 2 | 6.7% |
| Cronin et al. | 2 | 1 | 0 | 0 | 3 | 10.0% |
| Fritsch et al. | 5 | 0 | 0 | 0 | 5 | 16.7% |
| García-Mateo et al. | 2 | 0 | 0 | 0 | 2 | 6.7% |
| Haskey et al. | 3 | 0 | 0 | 0 | 3 | 10.0% |
| Lee et al. | 3 | 0 | 0 | 0 | 3 | 10.0% |
| Narimani et al. | 2 | 0 | 1 | 0 | 3 | 10.0% |
| Nikniaz et al. | 2 | 0 | 0 | 0 | 2 | 6.7% |
| Preda et al. | 3 | 0 | 1 | 0 | 4 | 13.3% |
| Seeger et al. | 1 | 0 | 0 | 0 | 1 | 3.3% |
| van de Pol et al. | 0 | 0 | 0 | 0 | 0 | 0.0% |
| **Subtotal** | **25** | **1** | **2** | **0** | **28** | **8.48%** |
| **Grand Total (57 studies, 1,710 cells)** | **90** | **4** | **46** | **11** | **151** | **8.83%** |

**Supplementary Table 5:** Qualitative characterization of expert modifications

| Evidence strength adjustment | Experts changed multiple statements from “should be considered” to “is appropriate” to match the RAND/UCLA formatting. And AI-statements were upgraded to a stronger recommendation. |
| --- | --- |
| Population expansion | AI-generated statements often focused on “obesity” alone as the population, whereas the experts expanded this to “obesity or overweight” across multiple interventions statements. |
| Terminology standardization | Minor corrections included preferred clinical nomenclature (“IBD flares” to “IBD exacerbations”, “Biologics” to “Advanced therapies” ), standardized abbreviations (“GLP-1 receptor agonists, GLP-1R agonists” to “GLP-1 RA”). |
| Sub-statements addition | Two composite statements were split into individually ratable sub-items to expand the scope of the statements. |
| Clinical scope redefinition | One statement discussed timing advanced therapy prior to bariatric surgeries and was changed to recommending bariatric surgeries regardless of therapy timing or type. |

**Supplementary Figure 1:** Study-level distribution of flagged cells per study, by subtopic

|  | Bariatric Procedures (n=24) | GLP-1RA (n=15) | Pharm CVD (n=7) | Diet & lifestyle (n=11) | All domains (N=57) |
| --- | --- | --- | --- | --- | --- |
| 0 (perfect extraction) | 2 | 0 | 0 | 1 | 3 |
| 1 | 7 | 1 | 0 | 1 | 9 |
| 2 | 4 | 3 | 3 | 3 | 13 |
| 3 | 8 | 3 | 2 | 4 | 17 |
| 4 | 3 | 5 | 1 | 1 | 10 |
| 5 (maximum observed) | 0 | 3 | 1 | 1 | 5 |
